# Patients with CLL have similar high risk of death upon the omicron variant of COVID-19 as previously during the pandemic

**DOI:** 10.1101/2022.03.01.22271685

**Authors:** Carsten U Niemann, Caspar da Cunha-Bang, Marie Helleberg, Sisse Rye Ostrowski, Christian Brieghel

## Abstract

Previous studies have shown that patients with chronic lymphocytic leukemia (CLL) and coronavirus disease 2019 (COVID-19) have high mortality rates. The omicron variant has been reported to give milder disease in the general population, but outcomes of infections with the omicron variant among immunocompromised patients have not previously been reported. In a population-based cohort we assessed rates of hospitalizations, ICU-admissions, and 30-day all-cause mortality among all patients with CLL from Eastern Denmark testing positive for severe adult respiratory syndrome coronavirus 2 (SARS-CoV-2) in time periods before and after dominance of the omicron variant. Rates of hospitalizations and ICU-admissions declined significantly, whereas 30-day mortality remained as high as 23% in the period with dominance of the omicron sublineage BA.2 variant. Thus, patients with CLL in general and in particular those above 70 years of age with one or more comorbidities should be considered for closer monitoring and pre-emptive antiviral therapy upon a positive SARS-CoV-2 test.

**Key points:**

- The omicron variant of COVID-19 leads to high fatality rates in CLL, despite milder disease in the background population
- Patients with CLL who test positive for SARS-CoV-2 in the era of the omicron variant should be considered for pre-emptive antiviral therapy

**Explanation of novelty:** The omicron variant has been reported to give milder disease in the general population, but outcomes of infections with the omicron variant among immunocompromised patients have not previously been reported. These population-based data on outcome for patients with CLL upon infection with the omicron variant of SARS-CoV-2 warrants closer monitoring and pre-emptive antiviral therapy upon a positive SARS-CoV-2 test for patients with CLL.

## Introduction

Patients with chronic lymphocytic leukemia (CLL) have increased morbidity and mortality following infection with severe adult respiratory syndrome coronavirus 2 (SARS-CoV-2) leading to coronavirus disease 2019 (COVID-19).^1,2^ The immune dysfunction inherent to CLL itself and CLL treatment whether targeted or chemoimmunotherapy based,^3^ is considered the likely cause of increased susceptibility to severe COVID-19. During the first and second pandemic wave, most CLL patients with COVID-19 developed severe disease and 30-day mortality was 31-50% for those admitted.^1,2^ Further, patients with CLL demonstrated impaired vaccination response in terms of ability to produce neutralizing anti-SARS-CoV-2 antibodies, while the T-cell response remained intact for more patients.^4-7^ Data on outcome upon infection with SARS-CoV-2 B.1.1.529 (omicron) variant is warranted for immunocompromised patients in general and for patients with CLL in particular.^8^ The first Danish omicron case was detected on 25^th^ Nov 2021, became dominant in Denmark by 17^th^ Dec 2021, enabling high levels of break-through infections among vaccinated individuals.^9^

In Denmark, all patients diagnosed with a hematological malignancy were offered third and fourth booster vaccination against SARS-CoV-2 in August 2021 and January 2022, respectively. At the same time, a single dose of anti-SARS-CoV-2 monoclonal antibody (mAbs) was recommended for immunocompromised patients testing positive for SARS-CoV-2, while the standard of care for immunocompromised patients admitted with moderate to severe COVID-19 was dexamethasone, heparin and remdesivir.^10,11^ Remdesivir was widely used for hematological patients regardless of disease severity since approval mid-2020.^12,13^ The mAbs sotrovimab retain its neutralizing activities against the omicron BA.1 sublineage, but very recently, in vitro studies have shown reduced activity against the BA.2 sublineage.^14 15^

## Methods

Insights into potential variation in clinical outcome for immunocompromised patients upon infection with the omicron variant is limited. Here we investigated the rate of hospitalization, admission to intensive care unit (ICU) and mortality following infection with SARS-CoV-2 among patients with CLL in a Danish population-based cohort from March 2020 through January 2022. As data on variants were missing for most patients, we grouped patients into four time periods based on the first positive SARS-CoV-2 PCR: Period 1: March 2020 - December 2020; Period 2: January 2021 - 25^th^ November 2021 (first omicron case in Denmark); Period 3: 26^th^ November 2021 - 31^st^ December 2021; Period 4: 1^st^ January 2022 - 28^th^ January (omicron variant dominating from 17^th^ December 2021 and sublineage BA.2 dominating from 1^st^ January 2022). Data were retrieved from electronic health records (EHR) covering a background population of approximately 2.8 million individuals.^16^ We included all patients registered with a CLL diagnosis (DC91.1) and a positive PCR for SARS-CoV-2 within the EHR. Patients with multiple positive PCR tests within 12 weeks were considered as having one infection. Baseline characteristics were stratified by time-period of first positive SARS-CoV-2 PCR test (Table 1). Primary outcomes were time to hospital admission, time to ICU admission and 30-day mortality. We followed patients from date of first positive PCR until event, death or date of last follow-up (22^nd^ Feb 2022). The study was approved by the Ethics Committee and Data Protection Agency.

**Table 1:** Characteristics for patients, based on time intervals of COVID-19 positive test; Period 1: March 2020 to 31/12-2020; Period 2: 1/1-2021 to 25/11-2021, Period 3: 26/11-2021 to 31/12-2021; Period 4: 1/1-2022 to 28/1-2022. ALC: Absolute Lymphocyte Count and Hemoglobin levels are reported at time of positive SARS-CoV-2 test (within 5 days of first positive test). Percentage and numbers of patients receiving monoclonal antibodies against COVID (mAbs), remdesivir and dexamethasone treatment upon COVID for the different time periods are provided.

| Variable |  | Period 1 | Period 2 | Period 3 | Period 4 | Total |
| --- | --- | --- | --- | --- | --- | --- |
| Age | median [IQR] | 71<br>[64.5, 80.5] | 77<br>[68.2, 82.0] | 76<br>[72, 81] | 74<br>[69.5, 83.0] | 75<br>[66.5, 82.0] |
| Sex | Male (%) | 32 (54.2) | 23 (57.5) | 12 (57.1) | 20 (64.5) | 87 (57.6) |
| IGHV | Mutated (%) | 16 (59.3) | 15 (60.0) | 12 (66.7) | 13 (59.1) | 56 (60.9) |
|  | missing | 32 | 15 | 3 | 10 | 60 |
| FISH | del(13q) (%) | 14 (51.9) | 8 (30.8) | 10 (50.0) | 13 (56.5) | 45 (46.9) |
|  | Normal (%) | 7 (25.9) | 9 (34.6) | 4 (20.0) | 1 (4.3) | 21 (21.9) |
|  | tri(12) (%) | 3 (11.1) | 3 (11.5) | 2 (10.0) | 5 (21.7) | 13 (13.5) |
|  | del(11q) (%) | 1 (3.7) | 3 (11.5) | 3 (15.0) | 2 (8.7) | 9 (9.4) |
|  | del(17p) (%) | 2 (7.4) | 3 (11.5) | 1 (5.0) | 2 (8.7) | 8 (8.3) |
|  | missing | 32 | 14 | 1 | 8 | 55 |
| Hemoglobin<br>g/dL | median<br>[IQR] | 10.6<br>[9.6, 11.9] | 11.9<br>[11.3, 13.0] | 10.6 [9.1, 11.5] | 11.4<br>[10.0, 13.3] | 11.4<br>[9.9, 12.7] |
|  | missing | 32 | 16 | 13 | 20 | 81 |
| ALC, 109/L | median<br>[IQR] | 2.7<br>[1.1, 6.9] | 6.1<br>[1.5, 27.0] | 8.3<br>[0.6, 77.2] | 1.2<br>[0.6, 10.8] | 3.2<br>[1.0, 12.6] |
|  | missing | 42 | 21 | 16 | 21 | 100 |
| Dexamethasone | Received (%) | 22 (37.9) | 21 (53.8) | 18 (56.2) | 9 (40.9) | 70 (46.4) |
| Remdesivir | Received (%) | 13 (22.4) | 17 (43.6) | 12 (37.5) | 4 (18.2) | 46 (30.5) |
| mAbs | Received (%) | 0 | 10 (25.6) | 15 (46.9) | 9 (40.9) | 34 (22.5) |

## Results and Discussion

Until 28^th^ January 2022, 151 patients with CLL were registered with 153 first positive PCR test for SARS-CoV-2 in the EHR system for Eastern Denmark, two patients with positive PCR tests more than a year apart were counted as two separate infections. There were no significant differences in baseline characteristics between the four periods (Table 1), at least 85 (56%) patients had received CLL therapy prior to testing positive for SARS-CoV-2. The rate of hospitalizations for patients with CLL testing positive for SARS-CoV-2 was highest (above 75%) during the second period, where preemptive mAbs were administered during hospital admissions for patients with CLL upon a positive SARS-CoV-2 PCR test (Figure 1A). During period 3 (omicron emergence) and period 4 (omicron dominance), mAbs were administered on an outpatient basis, which may in part explain the lower 30-day admission rates (56-60% vs 83%). ICU admission rates were highest prior to emergence of omicron (12-12.5% vs 0-3%, Figure 1B), which may reflect impact of a third COVID-19 vaccine and improved care for patients with COVID-19 as well as differences in severity between SARS-CoV-2 variants.^10,11^ The ICU admission rates in general were lower than previously reported in international cohorts of COVID-19 in CLL (26% to 37% for hospitalized patients).^1,2^ This could be due to the full implementation of early treatment with mAbs, almost universal treatment with remdesivir for hospitalized patients without renal failure and high vaccination rates.

**Fig 1:**
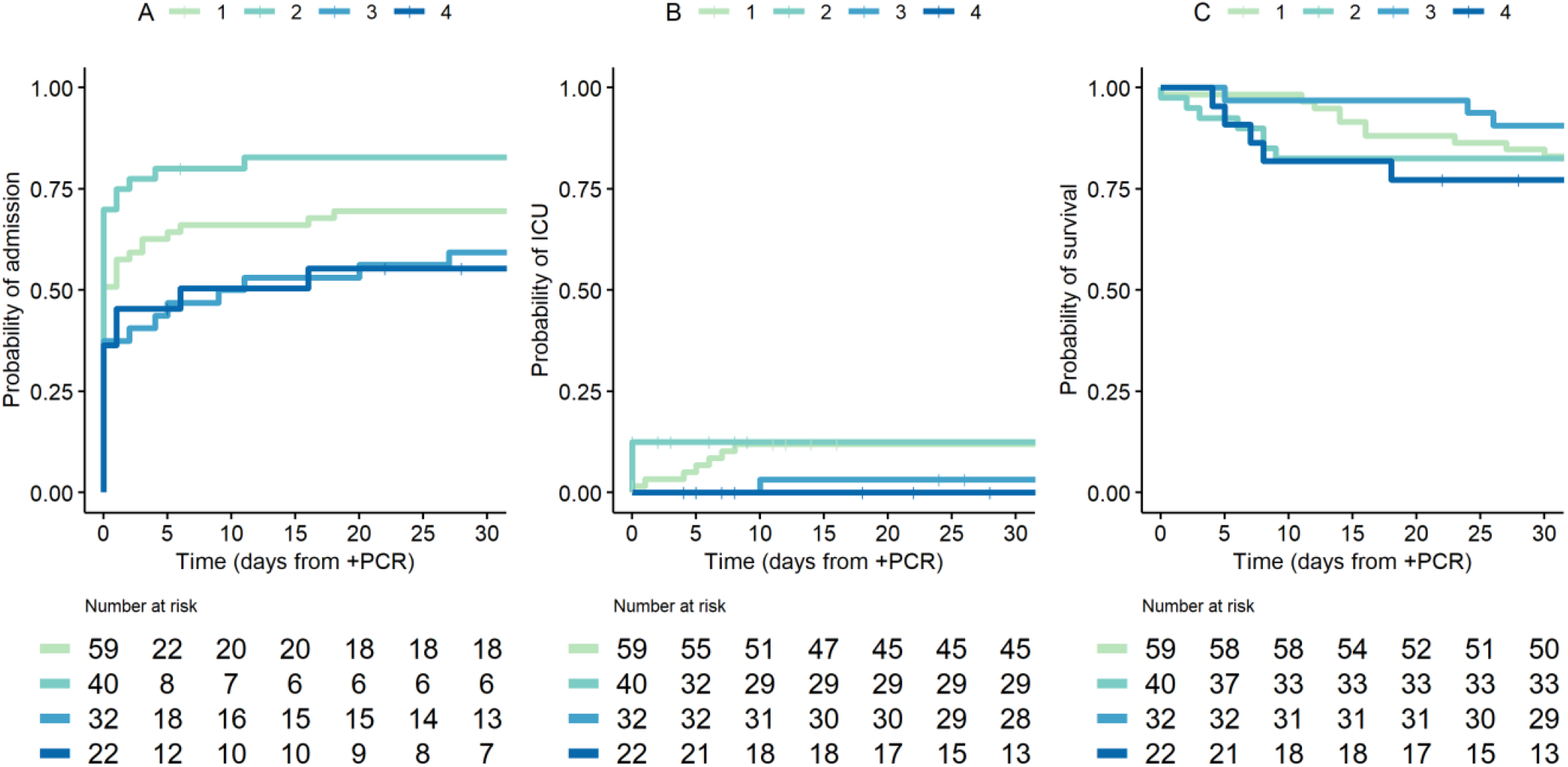
Kaplan-Meier curves for admission to hospital, admission to intensive care unit (ICU) and overall survival, for different time periods. Period 1: March 2020 to 31/12-2020; Period 2: 1/1-2021 to 25/11-2021, Period 3: 26/11-2021 to 31/12-2021; Period 4: 1/1-2022 to 28/1-2022.

Overall survival (OS) within 30 days after a positive PCR test for SARS-CoV-2 were above 75% for this immunocompromised population in all four time periods (77-91%). The estimated 30-day OS rate was lowest in period 4 when the omicron BA.2 sublineage was dominant, which may not be susceptible to Sotrovimab pre-emptive treatment.^15^ These survival rates are slightly better than the previously reported OS rates for COVID-19 in CLL during the first part of the pandemic (64-73%); although one study reported a higher OS rate of 89% for CLL patients testing positive for SARS-CoV-2 after 1^st^ May 2020.^1,2^ The estimated 30-day OS rate for period 3 seemed to indicate improved OS for CLL patients contracting COVID-19 after the emergence of the omicron variant. However, following patients for up to 90 days, the estimated OS rate decreased towards 75% for the first three periods with longer follow up. Five out of six fatal cases (including deaths after 30 days) in period 3 were the delta variant (missing variant information for the last case, data not shown). Patients positive for SARS-CoV-2 in period 4 (dominance of omicron sublineage BA.2) appeared to have a worse estimated 30-day OS rate (Figure 1C), while also receiving less COVID specific treatment (Table 1). The five patients who died within 30 days of a positive SARS-CoV-2 test in period 4 were aged above 71 years and all had comorbidities, e.g. dementia, other malignant diseases, diabetes, cardiac and pulmonary comorbidities. Four of the five fatal cases had confirmed omicron variant, while variant data were missing for the last. Three of the five patients died from respiratory failure while two patients died at home without known cause of death. Two of the fatal cases received mAbs and dexamethasone, one of them also remdesivir; the three remaining fatal cases did not receive COVID specific treatment.

Limitations apply to this study; the size of the patient population was limited, patients with CLL testing positive for SARS-CoV-2 at test sites not reported within the EHR may represent less severe CLL, thus causing overrepresentation of patients with more aggressive CLL in this study.

Based on epidemiological data from South Africa,^17^ the incidence of SARS-CoV-2 seems decoupled from the incidences of hospitalization and death upon emergence of the omicron variant, while previous vaccination seems not to protect against infection with the omicron variant of SARS-CoV-2. This study indicate that omicron sublineage BA.2 pose a similar risk of fatal COVID-19 for patients with impaired immune function due to CLL and/or treatment,^3,18^ with an estimated 30-day OS rate of 77%, as prior variants. Thus, patients above 70 with CLL and one or more comorbidities should be considered for closer monitoring and pre-emptive antiviral therapy upon a positive SARS-CoV-2 test.

## Data Availability

All data produced in the present study are available upon reasonable request to the authors with the restrictions that data privacy regulations put on the data set.

## Acknowledgement

The study was supported by a COVID-19 grant from the Ministry of Higher Education and Science (0238-00006B) and the Danish National Research Foundation (DNRF126); by the Danish Cancer Society and the EU funded CLL-CLUE for CUN. CB received funding from Weimann’s Legat. The Capital Region of Denmark, Center for Economy, provided data extracts from the EHR system.

## Authorship Contributions

SRO, CUN, CB and CdC developed the concept of the study, CB, CUN and CdC collected and curated data, CB performed statistical analyses, MH provided infectious disease and clinical perspectives and interpretation, CUN wrote the first draft of the paper, all authors contributed to and approved the final version

## Conflicts of interest

CUN received research funding and/or consultancy fees outside this work from Abbvie, Janssen, AstraZeneca, Beigene, Roche, CSL Behring, Takeda and Octapharma. CB received consultancy fess outside of this work from AstraZeneca. The remaining authors declare no conflicts of interest.

## Notes

### Author Declarations

The Ethics Committee of the Capital Region of Denmark gave ehtical approval for this work, Journal-nr.: H-20026502.

